## Supplemental Figures for "A Standardized Metric to Enhance Clinical Trial Design and Outcome Interpretation in Type 1 Diabetes"

[†]Authors contributed equally.

‡Authors contributed equally.

**Supplementary Figures:**

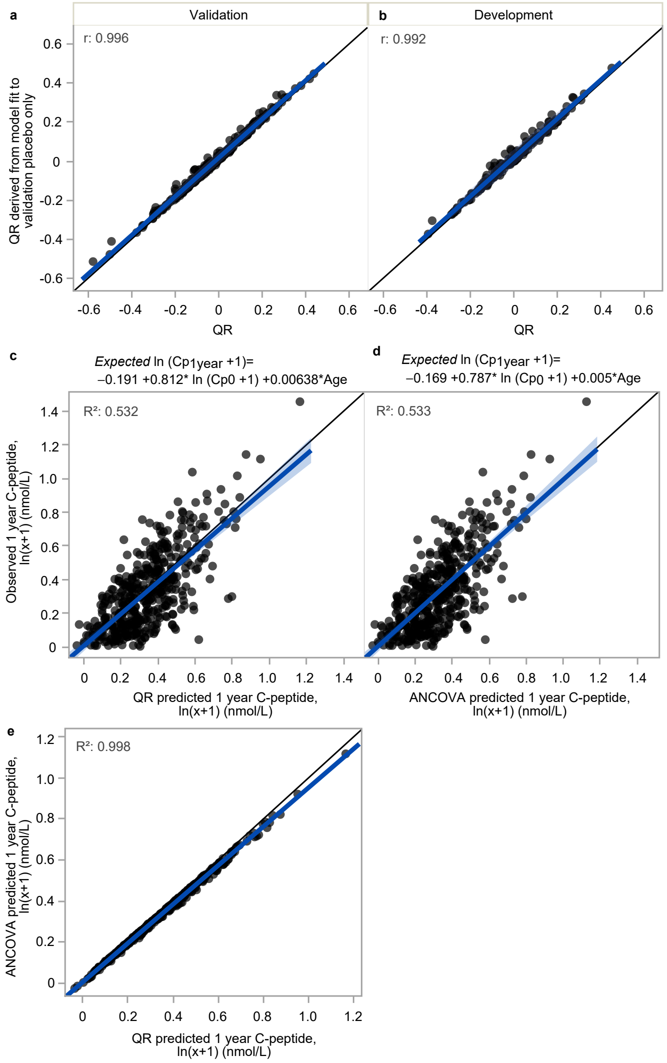


**Supplemental Figure 1. No difference between original and revised ANCOVA model to determine QR.**

QR derived from ANCOVA fit to validation cohort placebo/control participants only is strongly correlated with published QR model (i.e., model derived from development cohort) within both (**a**) validation (n=286) and (**b**) development (n=162) cohorts. Original ANCOVA model from 5 studies used in development of original QR model (**c**) compared with revised ANCOVA model using placebo/control participants from all 13 studies, n=448 (**d**). Predictive values between two models are strongly associated ($R^2$ =0.998) (**e**). Blue line indicates linear regression lines and shaded blue regions are 95% confidence intervals.

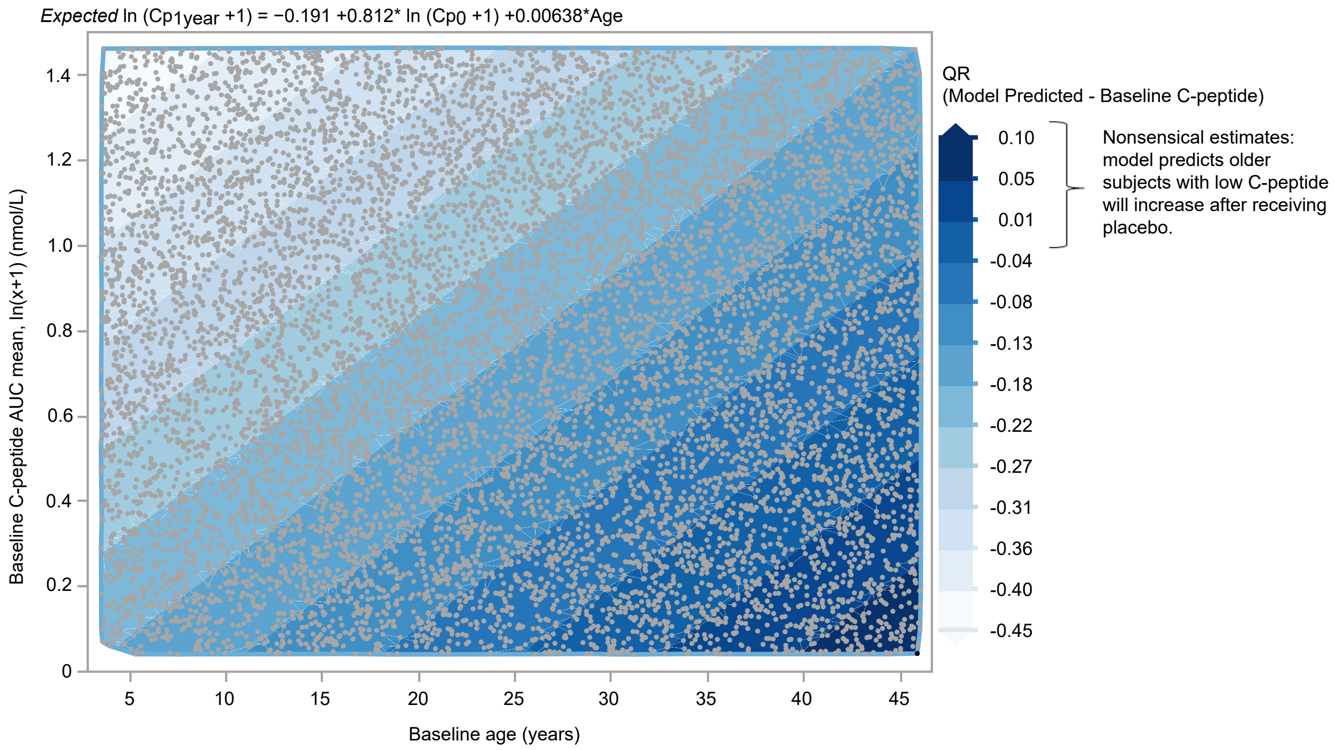


**Supplemental Figure 2. Evaluation of model predicted C-peptide across wide range of baseline C-peptide values and ages.**

Contour plot of model predicted C-peptide subtracted from baseline C-peptide across wide range of ages and baseline C-peptide. Dark blue regions in the lower right quadrant of the figure correspond to older subjects with low levels of baseline C-peptide where the model predicts subjects to have no decline or minor positive increase in C-peptide at 1 year.

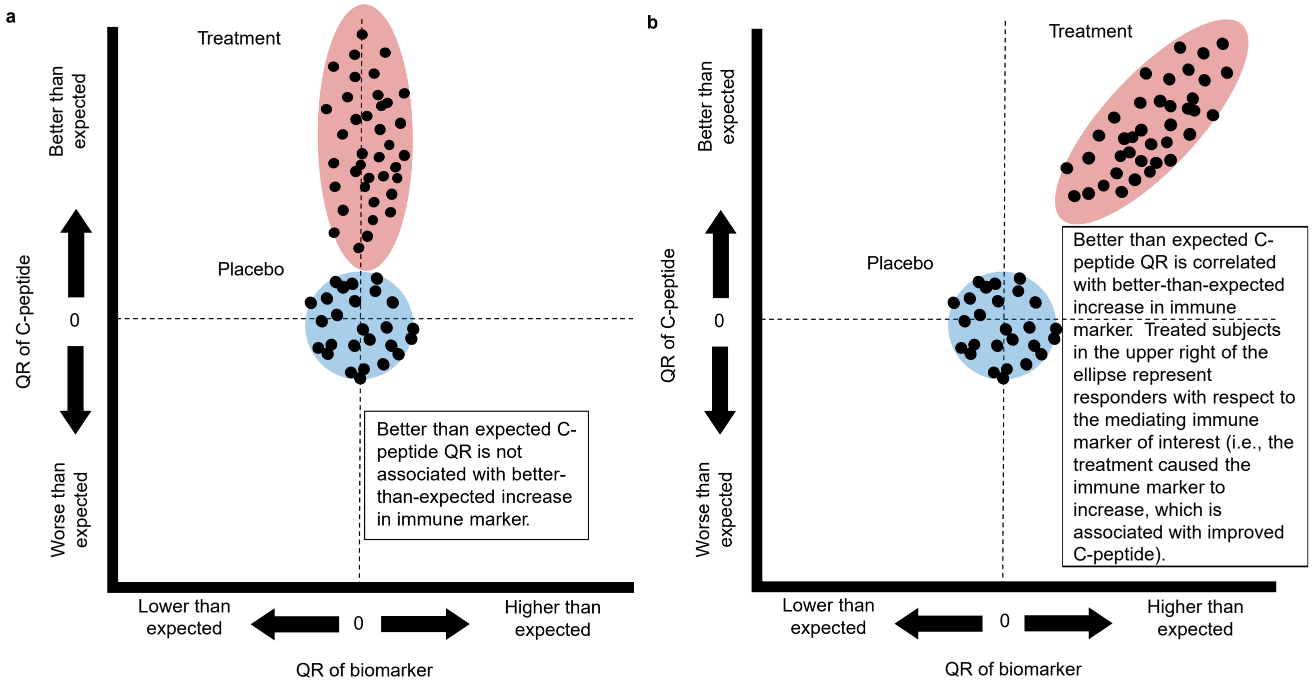


**Supplemental Figure 3. Multivariable adjustment with QR allows for assessment of causal association between biomarker and C-peptide.**

(**a**) Treatment is effective, but not associated with immune marker. (**b**) Evidence of causal association: better C-peptide response is correlated with biomarker among treated individuals.

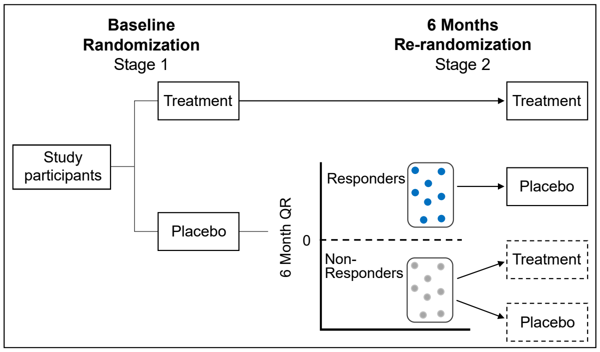


**Supplemental Figure 4. Sequential parallel comparison design (SPCD) to improve trial efficiency.**

SPCD re-randomizes placebo non-responders at 6 months and provides opportunity for additional individuals to potentially benefit from treatment as well as increasing efficiency of trial.
